# SARS-CoV-2 outbreak in a tri-national urban area is dominated by a B.1 lineage variant linked to mass gathering events

**DOI:** 10.1101/2020.09.01.20186155

**Authors:** Madlen Stange, Alfredo Mari, Tim Roloff, Helena MB Seth-Smith, Michael Schweitzer, Myrta Brunner, Karoline Leuzinger, Kirstine K. Søgaard, Alexander Gensch, Sarah Tschudin-Sutter, Simon Fuchs, Julia Bielicki, Hans Pargger, Martin Siegemund, Christian H Nickel, Roland Bingisser, Michael Osthoff, Stefano Bassetti, Rita Schneider-Sliwa, Manuel Battegay, Hans H Hirsch, Adrian Egli

## Abstract

**Background:** The first case of SARS-CoV-2 in Basel, Switzerland was detected on February 26^th^ 2020. We present a phylogenetic study to explore viral introduction and evolution during the exponential early phase of the local COVID-19 outbreak from February 26^th^ until March 23^rd^.

**Methods:** We sequenced SARS-CoV-2 naso-oropharyngeal swabs from positive 746 tests that were performed at the University Hospital Basel in the timeframe of our study. We successfully generated 468 high quality genomes from unique patients and called variants with our COVID-19 Pipeline (COVGAP). We analysed viral genetic diversity using PANGOLIN taxonomic lineages. To identify introduction and dissemination events we incorporated global SARS-CoV-2 genomes and inferred a time-calibrated phylogeny. We used epidemiological data to aid interpretation of phylogenetic patterns.

**Findings:** The early outbreak in Basel was dominated by lineage B.1 (83·6%), detected from March 2^nd^, although the first lineage identified was B.1.1. Within B.1, a clade defined by the SNP C15324T contains 68·2% of our samples (‘Basel cluster’), including 157 identical sequences at the root of the ‘Basel cluster’, suggesting local spreading events. We infer the origin of the ‘Basel cluster’ defining mutation to mid-February in our tri-national region. The remaining genomes map broadly over the global phylogenetic tree, evidencing several events of introduction from and/or dissemination to other regions of the world via travellers. We also observe family transmission events.

**Interpretation:** A single lineage variant dominated the outbreak in the City of Basel while other lineages such as the first (B1.1) did not propagate. We identify mass gathering events and less so travel returners and family transmission as causes for the local outbreak. We highlight the importance of adding specific questions to the epidemiological questionnaires that are collected, to obtain data on attendance of large gathering events and locations as well as travel history to effectively identify routes of transmissions in up-coming outbreaks. This phylogenetic analysis enriches epidemiological and contact tracing data, allowing, even retrospectively, connection of seemingly unconnected events, and can inform public health interventions.

## Introduction

The COVID-19 pandemic has rapidly spread around the globe during the first six months of 2020. The causative coronavirus, SARS-CoV-2, is the subject of many studies using genomic analysis providing key insights into viral diversity across cities^1^, provinces^2-5^, countries^6-11^, and globally^12^. SARS-CoV-2 has an estimated mutation rate of 0.71-1.40×10^−3 13^, which translates to 21-42 mutations per year. Due to the accumulation of mutations, phylogenetic analysis of SARS-CoV-2 is becoming more granular over time^14^, providing increasing resolution of transmission dynamics and events. Comparisons of single nucleotide polymorphisms (SNPs) allows us to explore transmission events with highest resolution across communities. The identification of transmission routes is important, especially with various public health measures being introduced, such as lockdown policies, which have been implemented on country or regional levels to limit viral transmission. The impact of public health measures can be monitored through phylogenies^3^. Genomic data can also deliver insights into mutations and whether they alter virulence, or aid adaptation to novel hosts. The spike protein D614G mutation, for example, has been implicated in more effective transmission^15^, although the actual impact may be through fixation in an expanding lineage rather than conferring increased transmissibility per se^16^.

The scope of this study is to provide a more granular picture of the phylogenetic diversification and propagation of the early-stage SARS-CoV-2 pandemic on a local scale. The City of Basel has a population of 175,350 inhabitants (median over the past five years) with half a million people in the Basel area. Situated in North-Western Switzerland, directly bordering both Germany and France, Basel has almost 34,000 workers commuting daily across the international borders^17^. Given this large exchange of people in this tri-national region, the fact that the neighbouring region Alsace, France, was already experiencing an intense epidemic^18^, and a low threshold testing strategy implemented weeks before the first case, we aim to explore the early stage of SARS-CoV-2 transmission dynamics in Basel and the surrounding area from the first case to one week post border closure.

## Results

### Characteristics of the longitudinal study

This cohort study includes all patient samples from Basel-City and the surrounding area during the initial 26 days of the local outbreak, between February 26^th^ and March 23^rd^. Only single, non-repeated tests per patient were considered eligible for phylogenetic analysis. This timeframe covers the first two positively tested cases in Basel on February 26^th^, which we were able to capture via early implementation of PCR-based detection by routine diagnostics, until the date of border closure plus seven days (March 23^rd^).

From the first case on February 26^th^ 2020 until March 23^rd^ we had performed 6,943 PCR tests. Of these, 746 samples (10.7%) were SARS-CoV-2 positive (**Figure 1**). March 23^rd^ had the maximum number of positive tests, with 66 cases. Of all PCR tested patients and positively tested patients during the study period, a majority were female with a median age of 42 and 49 years, respectively (**Table 1**). Of the PCR-confirmed cases, only 17 (2.3%) were in patients younger than 18 years. (**Table 1, Figure S1**). 418 (56%) were living in the canton of Basel-City (City of Basel, Riehen, Bettingen) and 328 (44%) were from the surrounding area.

**Table 1.**
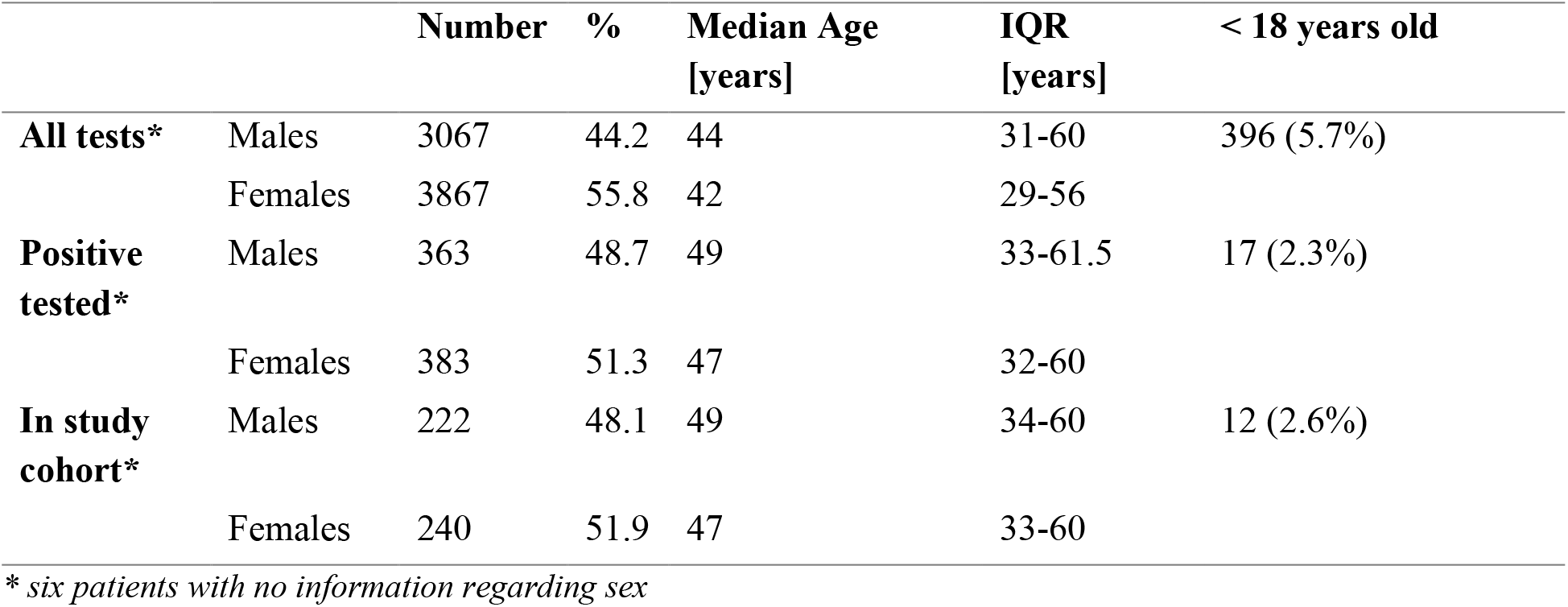
Number and age summary of all tested patients, positively tested patients, and patients with successfully sequenced SARS-CoV-2 genomes, by sex.

|  |  | Number | % | Median Age<br>[years] | IQR<br>[years] | < 18 years old |
| --- | --- | --- | --- | --- | --- | --- |
| **All tests\*** | Males | 3067 | 44.2 | 44 | 31-60 | 396 (5.7%) |
|  | Females | 3867 | 55.8 | 42 | 29-56 |  |
| **Positive tested\*** | Males | 363 | 48.7 | 49 | 33-61.5 | 17 (2.3%) |
|  | Females | 383 | 51.3 | 47 | 32-60 |  |
| **In study cohort\*** | Males | 222 | 48.1 | 49 | 34-60 | 12 (2.6%) |
|  | Females | 240 | 51.9 | 47 | 33-60 |  |

\* six patients with no information regarding sex

**Figure 1.**
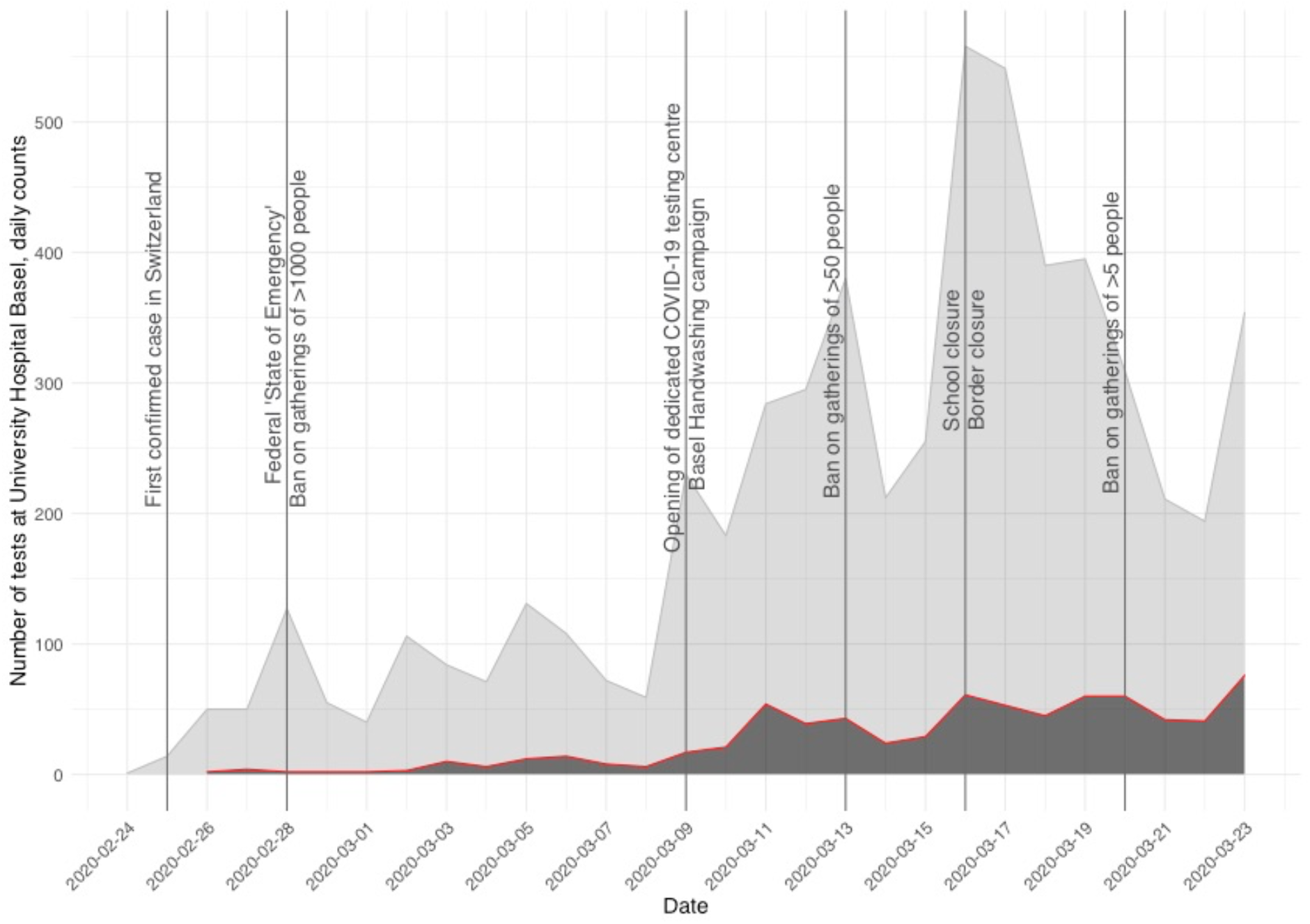
Epidemiological curve of the first COVID-19 wave in the city of Basel and hinterland, Switzerland. Positive (red line, dark grey area) and negative (light grey area) SARS-CoV-2 PCR tests are depicted from the beginning of the outbreak in February to March 23, 2020. Major events and imposed restrictions are marked by horizontal lines. First confirmed cases in Switzerland and Basel were on February 25^th^ and February 26^th^, respectively.

### COVGAP pipeline validation

No false positive variants were called (**Figure S4, Table 2, Table S1**). Ambiguous mapping was responsible for the failure to call the three indels, resulting in insufficient (<70%) coverage to be reliably called as a variant. This validation allowed us to determine the specificity (100%), sensitivity (94.2%), and accuracy (100%) of COVGAP, thus confirming its accuracy in SNP detection, as well as its ability to call the majority of indels from short read data.

**Table 2.** Sensitivity, specificity, and accuracy of COVGAP.

|  | MOCK POSITIVE | MOCK NEGATIVE |
| --- | --- | --- |
| **COVGAP POSITIVE** | TP=180 | FN=11 |
| **COVGAP NEGATIVE** | FP=0 | TN=2541564 |

TP: true positive, FP: false positive, FN: false negative. Numbers represent cumulative counts of bases that were or were not mutated over the 16 test genomes.

The COVGAP pipeline produced 468 (63%) high quality genomes for subsequent analysis, of the 746 samples taken. The remaining samples were either not available (N = 57), did not pass sequencing quality control (N = 156), or were duplicates from the same patient (N = 65). These 468 samples are subsequently referred to as the Basel area cohort. Of these, 240 (51.9%) were from female patients (**Table 1, Figure S1**), and 12 (2.6%) were from patients younger than 18 years.

### Phylogenetic lineages observed over time in the Basel area cohort

Over the 26-day study period, 13 out of 91 globally circulating phylogenetic lineages were recorded in the Basel area; only one additional lineage was recorded in all Swiss sequences.

Lineage B.1 dominated the cases during the initial phase of the outbreak (**Figure 2**), with 83.6% (N = 391) of sequenced samples (**Table 3**), being recorded for the first time in Basel on March 2^nd^. The first patient diagnosed at our hospital, on February 26^th^, had a virus belonging to lineage B.1.1. This lineage is seen sporadically through the outbreak with a maximum of six sequenced cases from March 16^th^. Lineage B.1 is associated with the Italian outbreak^14^, yet both B.1.1 (35.7%) and B.1 (51.0%) were the most prevalent lineages in Italy during this time span (**Figure 3**). From March 13^th^, rarer lineages are seen in the Basel area, such as B.1.1.6, a lineage that is associated with an Austrian origin^14^. Only two cases from the A lineage and sub-lineages were sequenced in the Basel area cohort (**Table 3**).

**Table 3.** Number of cases harbouring the S-D614G mutation in spike protein encoding gene in each phylogenetic lineage (PANGOLIN definition ver. May 19) and total count, in Basel area cohort by March 23rd 2020.

| Phylogenetic lineage | Number of samples S<sup>G614</sup><br>(derived) | Number of samples S<sup>D614</sup><br>(ancestral) | Total counts |
| --- | --- | --- | --- |
| A.2 | 0 | 1 | 1 |
| A.5 | 0 | 1 | 1 |
| B | 2 | 6 | 8 |
| B.1 | 391 | 0 | 391 |
| B.1.1 | 36 | 0 | 36 |
| B.1.1.1 | 1 | 0 | 1 |
| B.1.1.10 | 2 | 0 | 2 |
| B.1.1.6 | 1 | 0 | 1 |
| B.1.5 | 12 | 0 | 12 |
| B.1.8 | 3 | 0 | 3 |
| B.10 | 0 | 1 | 1 |
| B.2 | 0 | 8 | 8 |
| B.2.1 | 0 | 2 | 2 |
| B.3 | 0 | 1 | 1 |
| Sum | **448** | **20** | **468** |

**Figure 2.**
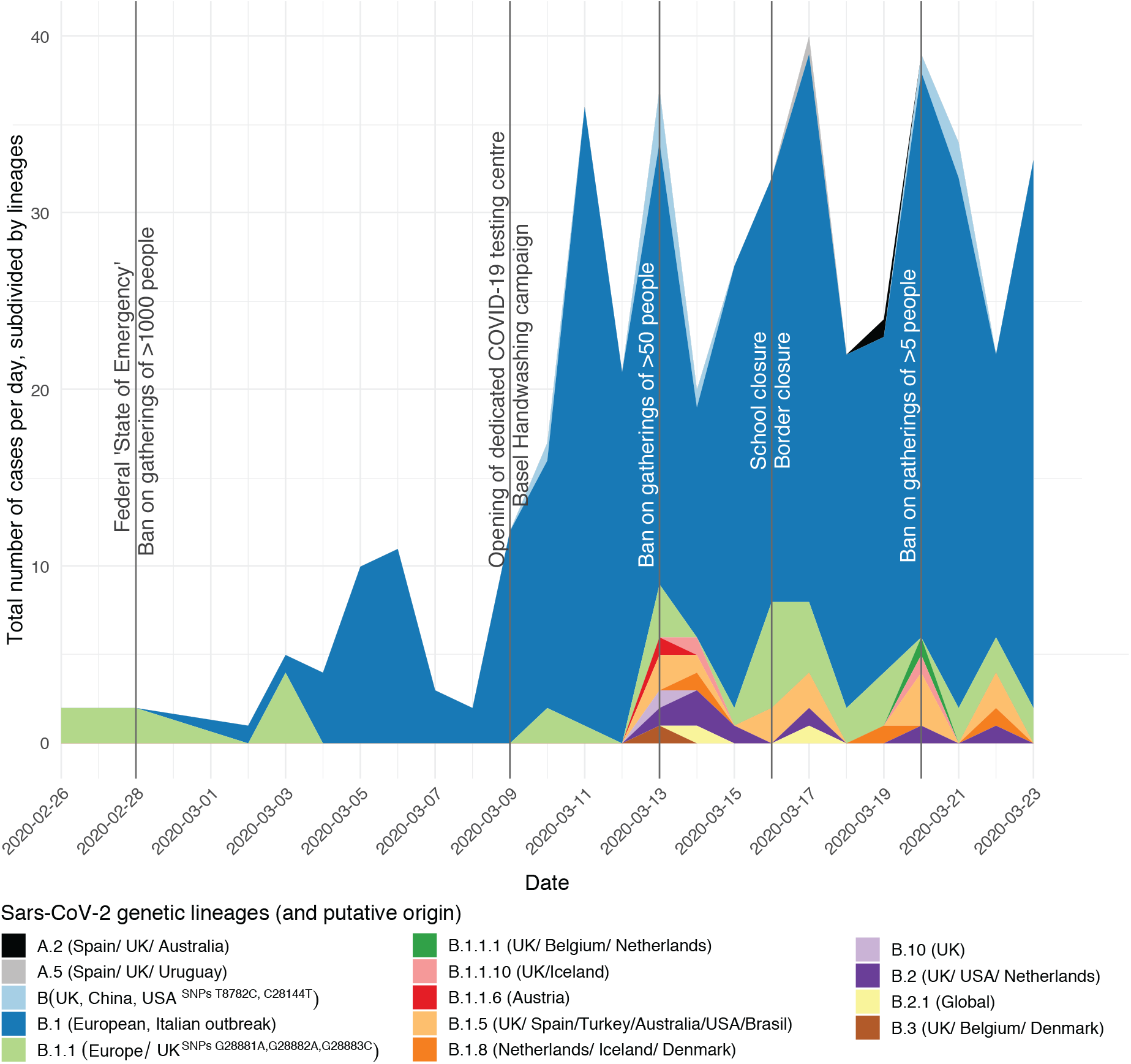
Detection of SARS-CoV-2 lineages found in the Basel area cohort from the first detected case on February 26^th^ to March 23^rd^ 2020. Major events and imposed restrictions are marked by horizontal lines. Low abundant lineages increase after the end of winter school vacation (March 8^th^) and are introduced by travel returners.

**Figure 3.**
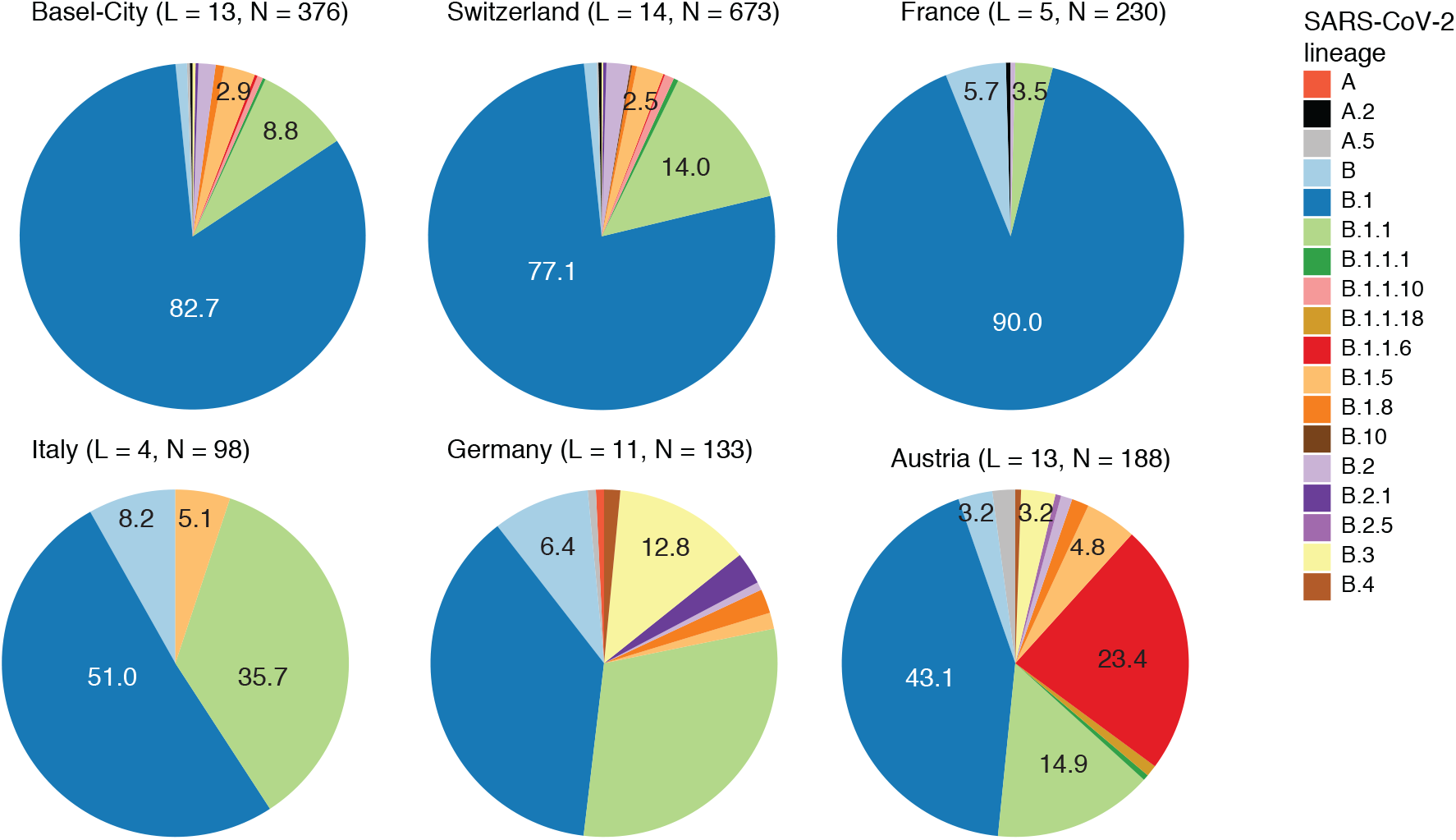
SARS-CoV-2 lineage diversity in neighbouring countries to Switzerland from first detected case until March 23^rd^, 2020. Number of lineages (L) and total number of genomes (N) per country in brackets, values within charts represent percentages. France was the first country in Europe that had confirmed COVID-19 cases on January 24^th^, followed by Germany on January 27^th^, Italy on January 31^st^, and some weeks later Austria and Switzerland followed on February 25^th^. Simpson diversity based on the available genomes and PANGOLIN lineage assignments is largest in Germany (3.87) and Austria (3.73), followed by Italy (2.52), it is smallest in Switzerland (1.62) and France (1.23). Basel-City mirrors the lineage proportions of Switzerland, while contributing half of Switzerland’s sequence data.

### Lineage diversity in Basel-City, Switzerland, and neighbouring countries

Comparison with lineages found in neighbouring countries over this period shows that lineage B.1 dominates in all, but the numbers of lineages identified, and the proportions vary (**Figure 3**). Switzerland (77.1%) has a similarly large proportion of B.1 lineage to France (90.0%). We describe the viral diversity based on abundance of lineages (retrieved from GISAID, for details see **Supplementary material)** using a range of diversity indices (**Table S5**). Simpson diversity, which accounts for differences in sample abundance between countries, was highest in Germany (3.87) and Austria (3.73), followed by Italy (2.52); it was lowest in Switzerland (1.62) and France (1.23). The share of our samples that originates from Basel-City residents (376 samples) excluding sequences that were obtained from commuters mirrors the lineage proportions of Switzerland (**Figure 3**), while contributing 56% of all available sequencing data for Switzerland within this timeframe (GISAID database as of June 22^nd^,^19,20^).

### Basel samples in global phylogenetic context

In order to better contextualize our findings, we analysed our virus genomes phylogenetically with a subset of global publicly available sequences (see **Supplementary material**; **Figure 4**). While phylogenetic lineages may show some geographical signal, lineages do not exclusively correspond to continents (**Figure 4A**), illustrating the degree of global interconnectivity and speed of spreading. The phylogenetic lineages recorded in the Basel area are distributed across the global phylogenetic tree (**Figure 4B**). Mismatch of ‘taxonomic’ assignment and phylogenetic origin of genomes assigned to B.1 can be seen, with several as yet unnamed sub-lineages apparent in the phylogeny (**Figure 4B**). We can identify a major clade, within lineage B.1, comprising 68.2% of our samples (319/468 samples with 264 (82.8%) from patients from cantons Basel-City and Basel-Landschaft; **Figure 5A**). The remaining Basel area sequences (31.8%) (**Figure 5 B-C, Figure S6 B-C**) are spread throughout the phylogeny and cluster with global genomes. Introductions and features of some of the clades are analysed in the following sections and in **Supplementary material**.

**Figure 4.**
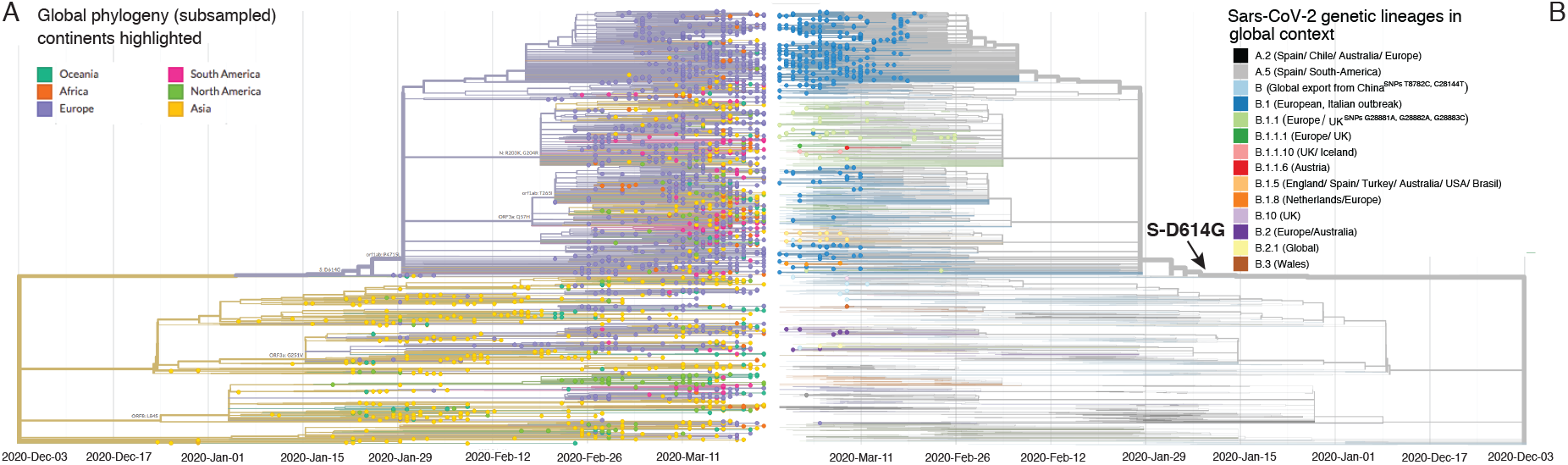
SARS-CoV-2 phylogeny of Basel area samples and genetic lineages (PANGOLIN) in a global context. **A**. Time tree of SARS-CoV-2 genomes from the Basel area cohort as well as subsampled global genomes (30 genomes per country and month), coloured by continent of origin.Amino acid mutations at internal nodes representing clade defining mutations are shown. **B**. Mirrored time tree coloured by genetic lineages sensu PANGOLIN v.May19 (https://github.com/cov-lineages/). Each tip with a circle represents a genome from the Basel area cohort, branches without circled tips represent global genomes, included to confer the global context of the Basel genomes.

**Figure 5.**
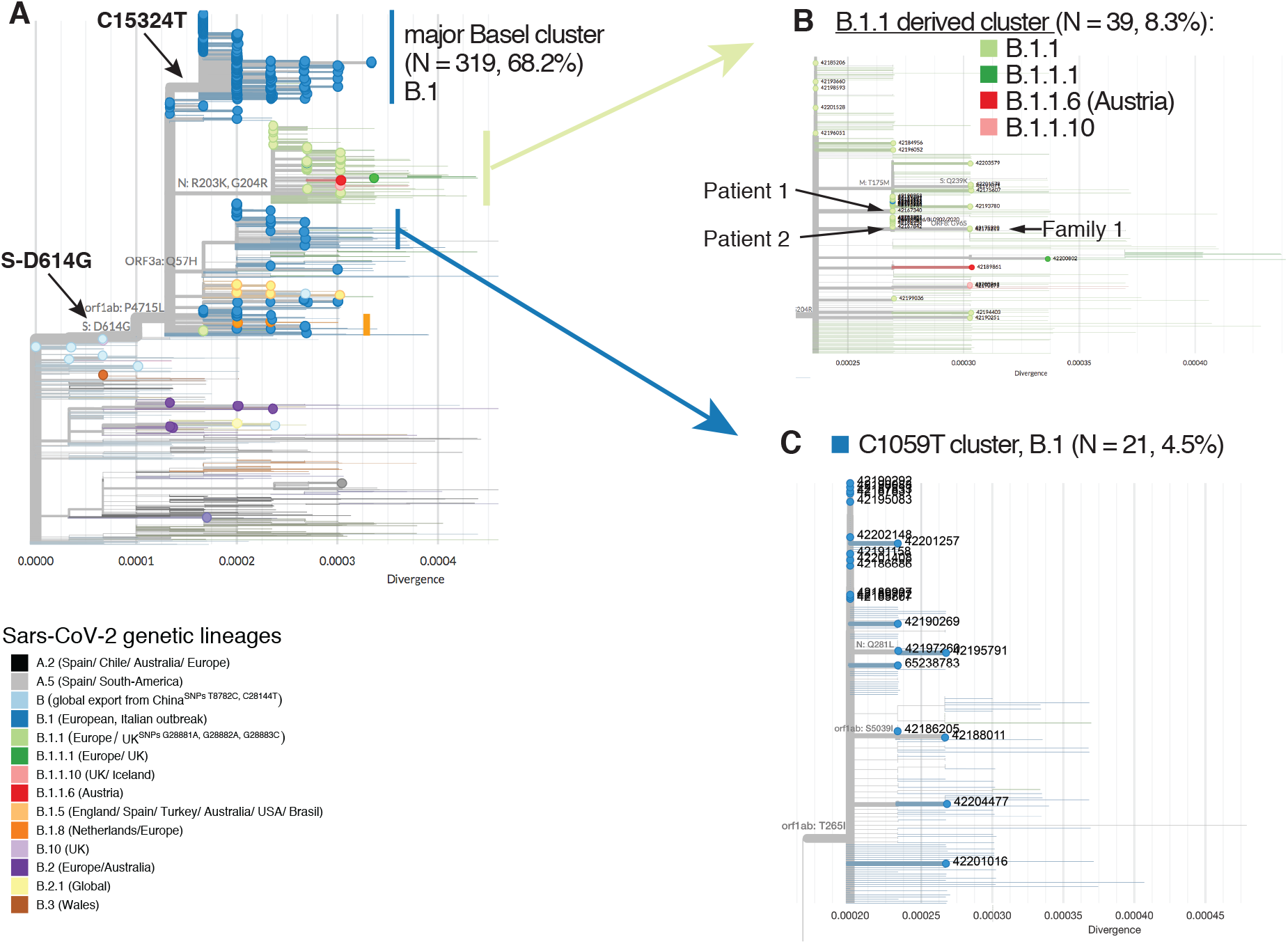
Divergence trees plotting nucleotide divergence between 468 genomes and expanded clusters of genomes in selected phylogenetic lineages. **A**. Genomes from Basel area cohort in global context. Tree composition is identical to the time tree from Figure 4. Branches with circles at the tip represent genomes from the present study; branches without circles represent global genomes from GISAID. The major Basel cluster contains samples with up to five mutations. **B**. Zoom into a mixed cluster derived from B.1.1 with seven to nine mutations difference to the root. A single genomes assigned to lineage B.1.1.6 has an assumed origin in Austria; two genomes (B.1.1.10) most likely originate from the UK. **C**. Potential ski-holiday related cluster (C1059T) with seven samples having a confirmed association with skiing destinations. Note: Divergence can be translated to number of mutations difference to the root (Wuhan-Hu-1) by multiplication by with the SARS-CoV-2 genome size (29903 bases).

### The first identified introduction of SARS-CoV-2 to Basel

The first two positively diagnosed patients with SARS-CoV-2 in Basel and first identified cases of COVID-19, *Patient 1* and *Patient 2*, travelled together to Italy. In our analysis, both *Patient 1* and *Patient 2* carried viruses from the B.1.1 lineage, the second most prevalent lineage in Italy at that time (**Figure 3**). Interestingly, the two virus genomes from *Patient 1* and *Patient 2* are separated by two SNPs, suggesting two independent infections (**Figure 5B**). Moreover, we did not identify minor alleles within these samples that would hint at double infection of the patients. In this case, the epidemiological cluster of *Patient 1* and *Patient 2* is not congruent with the phylogenetic inference. The virus genome of *Patient 1* carried a synonymous mutation at C313T in *ORF1ab*, which is found in samples from Israel, Hungary, Japan, USA, Argentina, Greece, India, Brazil, Morocco, and Netherlands among others (nextstrain.org), all sharing an unsampled common ancestor that emerged around February 25^th^ (CI February 23-26^th^). This SNP is also found in ten other Basel area cohort genomes: eight of these were from a family and social friends cluster unrelated to *Patient 1* that were diagnosed between March 13^th^ and March 22^nd^.

The virus genome of *Patient 2* carried a synonymous mutation at T19839C in *ORF1ab* and not C313T, and clusters together with eight identical virus samples sampled between February 26^th^ and March 23^rd^. Two of these are from family members of *Patient 2*, who tested positive two and six days later, suggesting a family route of infection. The epidemiological data of the other six patients suggests no direct transmission via *Patient 2*. One of the six returned from a Swiss ski resort three days prior to onset of symptoms. Two additional samples forming a family cluster (*Family 1*, **Figure 5B**) diagnosed on March 3^rd^, carried the T19839C plus non-synonymous mutation G28179A leading to amino acid change ORF8-G96S. One member of *Family 1* travelled with *Patients 1* and *2* to Italy and possibly got infected there with a yet different virus variant.

### Introduction of the Basel cluster

The clade within lineage B.1, into which 68.2% (N = 319) of our Basel area cohort sequences fall, is characterized by a synonymous SNP C15324T in *ORF1ab*, henceforth referred to as the “Basel cluster”. *Patient 1* and *Patient 2* are not linked to this Basel cluster. The first sample within the Basel cluster, from March 2^nd^, was from a patient residing in Central Switzerland, who was transferred to a care facility in Basel where six further people tested positive between March 5^th^ and March 17^th^ with identical viral genomes. The source of the first infection is unknown. The second sample within the Basel cluster was from a patient diagnosed on March 3^rd^, who had attended a religious event in Alsace, France, that took place between February 17^th^ and 21^st^. One other patient, who tested positive on March 9^th^ and had symptoms 14 days prior to testing, also attended this event.

### Description of the Basel cluster

The 319 genomes in the Basel cluster (Figure 5A) show a divergence of up to five SNPs up until March 23^rd^, although 157 genomes are identical and located at the root of the clade. This infers that the common ancestor originated between February 9^th^ and February 17^th^. That the clade defining SNP C15324T (GISAID emerging clades label 20A/15324T) was registered globally for the first time on March 2^nd^ simultaneously in the Basel area sample 42173111 and GISAID sample Germany/FrankfurtFFM7/2020 suggests unsampled circulation of this variant from mid-February. By searching all genomes available on GISAID (N = 80,189; as of August 12^th,^ 2020), filtering for genomes belonging emerging clade 20A/15324 and sampled until March 23^rd^ (N = 2,856), we find that subsequently the C15324T mutation has also been observed in other countries but remains most prevalent in Switzerland (NGISAID =57/213, 26.8%; NGISAID+this study = 386/675, 57.2%; first genome 42173111 from March 2^nd^) (**Table S3**). Of the 57 Swiss genomes with this mutation that were already deposited on GISAID, 26 also originate from the cantons Basel-City and Basel-Landschaft (Table S4). The mutation is also found in higher proportions in genomes from France (N = 69/369, 18.7%, first from March 3^rd^ sample France/HF1870/2020), Luxembourg (N = 24/116, 20.7%, from March 8^th^ sample Luxembourg/LNS2614631/2020), and Belgium (N = 40/268, 14.9%, from March 6^th^ sample Belgium/NKR-030645/2020), but date later than the first recorded occurrence from Switzerland and Germany (**Table S3**). Subsequently, as of March 9^th^, descendants of this variant were recorded outside Europe (**Table S4**) in smaller proportions than in Basel (**Table S3**), which suggests dissemination from the Basel area.

### Potential ski-holiday related cluster (C1059T)

Previous reports have identified viruses in lineage B.1 carrying SNP C1059T (amino acid change ORF1a-T265I) in travel returners from ski holidays in Ischgl, Austria^9,21^. We detected 21 (4.5%) viral samples within our cohort with these features (**Figure 5C**), divergent by up to two SNPs from the common ancestor. Samples date from March 11^th^ to March 23^rd^ with an inferred internal node age of February 21^st^ (CI: February 20^th^-February 21^st^, 2020), fitting with possible infections in Ischgl from end of February to March 13^th 22^. Epidemiological data confirms that five patients from that cluster had returned from Austria, and three specifically from Ischgl, the fourth from Tyrol, before testing positive. Two additional patients returned from skiing in Swiss ski resorts.

### Spike protein mutation prevalent in Basel patients

The spike protein S-D614G mutation is associated with the B.1 lineage and all those derived from this (**Figure 4**). As such, it occurs in 448 of the 468 (95.7%) samples from the Basel area. The SNP responsible has not been lost once in our sub-sampled dataset, but it is not present in B.2 or B.3 or other sister lineages to B.1 (**Table 3**). Among our samples, we found no significant difference in viral loads between patients with and without the S-D614G mutation (z = −0.881, p = 0.38). However, our cohort is biased to samples with higher viral load, as these were those that were successfully sequenced.

## Discussion

We reconstruct the early events focusing on introduction and spread of SARS-CoV-2 in Basel and the surrounding area, Switzerland, from a phylogenetic perspective. We present COVGAP, a new combination of existing tools to effectively and efficiently mine SARS-CoV-2 genomes from Illumina paired end reads. Unlike other currently available tools^23^, COVGAP shows higher sensitivity levels in SNP calling from raw reads (100%), failing only in ambiguously mapped deletions and insertions. In such cases, it adopts a coverage-conservative approach, needed to reliably call variants in real world scenarios.

The majority of genome variants in Basel are similar to those from France, Italy, and Germany. We found the presence of 13 SARS-CoV-2 lineages in our samples, with the beginning of the Basel outbreak being powered by the European B.1 lineage. In particular, a B.1 lineage variant with the C15324T mutation dominated the early phase of the local spread with 70% of samples forming a large Basel cluster. Compared to Victoria, Australia^5^, the UK^24^, or Austria (this study) the diversity seen arriving in Switzerland and Basel as determined using Simpson diversity is more limited, reflecting European rather than intercontinental connections. This diversity measure can be used to monitor viral introductions as an effect of travel restrictions in the future.

The Basel cluster virus variant 20A/C15324T was first detected in Europe on March 2^nd^ in Germany and Switzerland simultaneously. We locate its geographic origin to our tri-national region between February 9^th^ and February 17^th^. Our epidemiologically-informed phylogenetic analysis indicates that the Basel cluster represents a larger transmission chain that was unchecked and spread effectively among unrelated people throughout Basel and eventually outside of Europe. The first recognized case in the large Basel cluster goes back to a patient in a care facility, in which several more infections occurred.

The beginning of the COVID-19 outbreak described here coincided with the winter school holidays in Basel, from February 22^nd^ to March 8^th^. During this time, many residents take the opportunity to travel, in particular to skiing resorts. Viral introductions from ski resorts are known from contract tracing data to have affected Germany^25^, Denmark^21^, Iceland^9^, France, Spain, and UK^26^, with Ischgl, Austria being a described source of many cases. Our data supports the finding that skiing resorts in Austria and Switzerland served as dissemination hotspots. This school holiday also provided the opportunity for the first identified SARS-CoV-2 introductions to Basel through two jointly returned travellers, notably each with different viral variants. Overall, however travel returners did not drive the outbreak in Basel evidenced by the low diversity of variants and proportion of such variants in our sample. A second likely source represents the many workers travelling daily across borders from France and Germany, particularly from heavily affected areas such as Alsace^18^. As the B.1 and B.1.1 lineages were dominant in France and Germany, these may be some sources of cases and transmissions.

The timing of the epidemic in Basel also coincided with three major events. Firstly, a religious event from February 17^th^ to 21^st^ in Alsace that was described as a super-spreading event in France^27^. We confirmed that the virus genomes of two patients known to have attended are indeed situated at the root of the clade that constitutes the Basel outbreak. Secondly, carnival in the Basel area is celebrated over several weeks from early January, with numerous events, and thousands of active participants. The culmination is the UNESCO world-heritage ‘Basler Fasnacht’, scheduled this year (2020) for March 2^nd^-4^th^, but cancelled due to COVID-19. Notably, the active participants practice the piccolo and drums over weeks in closed rooms as a preparation for their performance during the carnival, and unofficial events are likely to have taken place. Thirdly, Basel hosted three international soccer events at the St. Jakob Stadium on February 15^th^ (20,675 spectators), 23^rd^ (20,265 spectators), and 27^th^ (14,428 spectators). All three major events fell around the inferred date of origin of the Basel mutation C15324T and subsequent dissemination phase.

A limitation of the current study is that we are likely to have missed some cases as not all symptomatic people were advised to be tested, especially children younger than 18 years old. Nevertheless, our cohort represents a very high sequencing density per detected case for a city (468 genomes from 746 PCR-confirmed cases in Basel area (62.7%) and from 10,680 PCR-confirmed cases nationwide (4.4%)) for this early phase of the pandemic^28^.

The availability and integration of epidemiological data in the interpretation of phylogenetic clades underlines the validity of instrumentalising those tools for improving the understanding of SARS-CoV-2 outbreak dynamics. Utilizing the clades as the backbone for targeted epidemiological analysis of specific cases helped in grasping how mass gatherings, travel returners, and care facilities may influence an outbreak within a city. The epidemiological data that was collected for the Federal Office of Public Health (FOPH), as requested by law, helped tremendously to verify travel related links; however it was not designed to obtain data on local super-spreading events such as attendance to soccer games, visiting clubs, restaurants, bars, and concerts and future versions could be improved. The classical epidemiological context is very important to further explain molecular epidemiological links especially in a still not very diversified virus.

In conclusion, the start of the outbreak of SARS-CoV-2 in the Basel area was characterized by a dominant variant, C15324T, within the B.1 lineage, which we infer to have arisen in mid-February in our tri-national region. Large gatherings (potential super spreading events) could have had profound effects on outbreak dynamics. Improved surveillance measures are needed in the management of an outbreak, including large-scale, active screening in the broader public, including more children to assess their role in transmission. Our analysis shows the potential of molecular epidemiology to support classical contact tracing, even retrospectively, in order to evaluate and improve measures to contain epidemics like COVID-19.

## Materials and Methods

### Patients, samples, and diagnosis

Respiratory samples from the University Hospital Basel and the University Children’s Hospital Basel (UKBB) patients were tested for SARS-CoV-2: from January 23^rd^ 2020 testing was based on current case definitions from the Federal Office of Public Health (FOPH); from 27^th^ February additionally, all respiratory samples negative for other respiratory pathogens were tested. Patient samples which tested positive for SARS-CoV-2 ^29,30^ up to and including March 23^rd^ were considered eligible for the present study. In total 6,943 diagnostic tests were performed during the study period. The 746 positively tested cases came predominantly from the administrative unit of Basel-City, Riehen, and Bettingen (418, 58%), while the remaining patients were from Basel-Landschaft and neighbouring cantons and countries.

For diagnosis, swabs from the naso- and oropharyngeal sites (NOPS) were taken, and combined into one universal transport medium tube (UTM, Copan). Total nucleic acids (TNAs) were extracted using the MagNA Pure 96 system and the DNA and viral RNA small volume kit (Roche Diagnostics, Rotkreuz, Switzerland) or using the Abbott m2000 Realtime System and the Abbott sample preparation system reagent kit (Abbott, Baar, Switzerland). Aliquots of extractions were sent for diagnosis to Charité, Berlin, Germany from January 23^rd^ - 29^th^, and to Geneva to the National Reference Centre (NAVI) in Switzerland from January 29^th^. In-house analysis started February 27^th^ as part of the hospital routine diagnostics as previously described ^30^.

### Whole genome sequencing (WGS)

SARS-CoV-2 genomes were amplified following the amplicon sequencing strategy of the ARTIC protocol (https://artic.network/ncov-2019) with V.1 or V.3 primers ^31^. In detail, real-time reverse transcriptase (RT) reactions were run to a total volume of 10 μl extracted total nucleic acid. After some optimization, PCR used 25 cycles for samples with a diagnostic cycle threshold (Ct) value lower than 21 (viral loads higher than 8.2 log10 Geq/ml); 40 cycles for all other samples (lower viral load samples) and repeats. Purified amplicons were converted into Illumina libraries with Nextera Flex DNA library prep kit (Illumina) automated on a Hamilton STAR robot, using 5ng input DNA. 96 libraries were multiplexed and sequenced paired-end 150 nucleotides on an Illumina NextSeq 500 instrument.

#### Consensus sequence generation and detection of mutations

After demultiplexing using bcl2fastq software version v.2.17 (Illumina), COVGAP (COVid-19 Genome Analysis Pipeline) was used (**Figure S2**). This incorporates: quality filtering using trimmomatic software version v.0.38 ^32^ to remove Illumina adaptors and PCR primer sequences from read ends; removal of reads smaller than 127 bases, and removal of reads with a phred score under 20 (calculated across a 4-base sliding window). Quality filtered reads were mapped to the Wuhan-Hu-1 reference MN908947.3 ^33^ using the BWA aligner ^34^. Reads flagged as mapping to the reference were retained ^35^, and are deposited under project PRJEB39887. SNPs and indels with respect to the reference sequence were called using pilon version 1.23 ^36^. Pilon summary metrics ‘alternative allele fraction’ (AF) and ‘depth of valid reads in pileup’ (DP) were used to identify major and minor alleles across all bases, which is not implemented in pilon itself. Major alleles were called if supported by 70% of the reads covering the variant locus (AF) for any locus with a minimum of 50x coverage (DP). Variants were applied to the reference to produce a consensus sequence; any base position with less than 50x coverage was masked with ambiguous characters (Ns) using BCFTools version 1.10.2 ^37^. Consensus sequences were accepted for further analysis when containing up to 10% Ns. Summary statistics, logs, coverage plots, and genome stack plots were generated using R version 3.6.0 and packages Gviz v1.30 ^38^, Sushi v1.23 ^39^, seqinr v3.6.1 ^40^, and ggplot2 v3.11 ^41^. COVGAP also provides per genome quality control visual outputs (**Figure S5**) and is available at https://github.com/appliedmicrobiologyresearch.

Quality control statistics such as the relationship between Ct-value and number of mapped reads and coverage are presented in **Figure S3**. In general, we observed a negative trend linking Ct values and percentage of ambiguous bases (Ns) being called as a result of low coverage. Sequences passing the quality filter (n=533) showed a lower Ct value (median: 22.4±5.14) than the ones that failed (n =156; median: 35.75.9±5.75).

#### COVGAP Validation

We used a set of 15 randomly *in silico* mutated SARS-CoV-2 mock genomes for the validation of the specificity (identification of true negatives) and accuracy (identification of true negatives and true positives) of COVGAP. Additionally, the genome MT339040, which harbours an 81 nucleotide deletion in the ORF7a gene and a further seven SNPs relative to the reference ^42^ was used. Together, the mock genomes possess 38 mutations including 30 SNPs, six deletions and two insertions across the reference genome MN908947.3. The genomes were then shredded to artificial paired-end 150 nucleotide reads using SAMtools wgsim ^37^ and processed by COVGAP. For validation purposes, original mock genomes and the COVGAP generated genomes from the shredded reads were aligned using Seaview v4.6 ^43^ and clustalw ^44^. A phylogeny was built using PhyML within Seaview with default parameters.

### Phylogenetic lineage assignment of Basel samples

To assess the phylogenetic diversity of SARS-CoV-2 samples during the early phase of the pandemic we inferred the lineage assignment for each consensus sequence derived from the COVGAP pipeline using PANGOLIN ver. May 19th (Phylogenetic Assignment of Named Global Outbreak LINeages) ^14^ available at github.com/hCoV-2019/pangolin. Details on lineage summaries, describing which countries lineages have been reported from and where transmission events have been recorded, can be found at https://github.com/hCoV-2019/lineages. Lineage assignments were used to aid visualization of phylogenetic diversity in Basel in a global context. For global sequences we used the PANGOLIN lineage assignments as provided by GISAID (https://www.gisaid.org/; ^19,20^) (details next section), which were used for plotting purposes on phylogenetic trees.

To compare the lineage diversity in Switzerland and Basel-City to neighbouring European countries (Austria, France, Germany, and Italy) during the early phase of the pandemic, we visualized relative abundances of lineages using all high-quality, on GISAID (downloaded June 22^nd^, 2020) available consensus sequences for the time until March 23^rd^ from Austria (N = 188), France (N = 230), Germany (N = 133), and Italy (N = 98). For Switzerland (N = 673), we combined our sequences (N = 468) with other sequence data from Switzerland ^48^ published on GISAID. To infer the diversity for canton Basel-City (including Bettingen and Riehen) excluding sequences that were obtained from commuters, we used the Basel-City portion (N = 376) of the Basel area cohort excluding samples from patients from cities outside of the administrative district of Basel-City. We calculated Simpson diversity (inverse Simpson concentration) as implemented in the SpadeR package v.0.0.1 ^45-47^, which controls for lineage abundance differences between the countries, which is dependent on available sequence data, and which ranges from 0 (no diversity) to indefinite (large diversity).

### Analysing Basel SARS-CoV-2 genomes in global phylogenetic context

High-quality and full-length consensus sequences and corresponding metadata (sample ID, date of sample, geographic location of sampling, PANGOLIN lineage) from Swiss ^48^ and global viruses were downloaded from GISAID on June 22^nd^, 2020, making 49,284 individual genome sequences. 43,252 sequences were retained after filtering for genomes with under 10% ambiguous characters (Ns) (author Genivaldo Gueiros Z. Silva) ^49^. Metadata and consensus sequences of the Basel samples and global data from GISAID were combined for further joint analysis, which were performed using custom R scripts and the nextstrain command line interface analysis pipeline v.2.0.0 (nextstrain.org) and augur v.8.0.0 ^50^.

Dates in our study samples correspond to date of sampling. Sequences were filtered by date from December 1^st^ 2019 to March 23^rd^ 2020 using an R custom script in R version 4.0.0 ^51^ and packages tidyr ver.1.1.0 ^52^, dplyr ver. 1.0.0. ^53^, and readr ver. 1.3.1. ^54^: 15,973 consensus sequences, including the Basel area sequences, remained. These time-filtered sequences were sub-sampled by geographic location to 30 sequences per country and month. Non-human derived viruses as well as sequences with other ambiguous characters (Us), as well as those from cruise ships, and duplicated sequences defined by the nextstrain team as of June 24^th^ (https://github.com/nextstrain/ncov/) were excluded using *augur filter* ^50^ resulting in 2,485 sequences for the final phylogenetic analysis dataset.

Consensus sequences were aligned to the NCBI Refseq sequence Wuhan-Hu-1 reference MN908947.3 using mafft v7.467 with method FFT-NS-fragment ^55^ and options --reorder –keeplength --mapout --kimura 1 -- addfragments --auto. The resulting alignment was end-trimmed to remove low-quality bases (bases 1-55; 29804-29903). We masked homoplasic sites (**Table S2**) that have no phylogenetic signal ^56^ (deposited at https://github.com/W-L/ProblematicSites_SARS-CoV2). Please note, that this list is under constant development as number and diversity of sequence data evolves; we retrieved the data on June 19^th^, 2020. Masking was done using *augur mask*.

The resulting alignment was analysed in IQ-TREE 2 ^57^ for tree inference using *augur tree* with substitution model GTR+G. The tree in Newick format was then subjected, together with the date information of each genome and the initial sequence alignment, to an estimation of the evolutionary rate by a regression of the divergence (number of mutations) against the sampling date using TreeTime ^58^ implemented in *augur refine*. Genomes or branches that deviated more than four interquartile ranges from the root to the tip versus the time tree were removed as likely outliers. The resulting time-calibrated and divergence trees were re-rooted to MN908947.3 and MT291826.1, the first official cases and published genomes of SARS-CoV-2 from Wuhan, China.

Ancestral trait reconstruction of each patient’s viral genome was done for region (continent) and country as well as region and country of exposure using *augur* traits with a sampling bias correction of 2.5. Internal nodes and tips (actual genomes) were annotated regarding their nucleotide and amino acid changes in relation to the reference using *augur ancestral* and *augur translate*, respectively. All data were exported as json files (supplementary files) using *augur export v2* to be visualized in *auspice* v2 ^50^.

Identified clades of interest were further inspected for existing epidemiological links using data collected by the University Hospital.

### Identifying genomes belonging to GISAID emerging clade A20/15324T

To identify a possible geographic origin of the synonymous C15324T mutation in *ORF1ab*, we performed a search on all available GISAID genomes as of August 12^th^, 2020. We downloaded all high quality and complete genomes that were assigned do GISAID legacy clade G (corresponds to clade 20A) and PANGOLIN lineage B.1 (all three are mostly congruent ^59^) with a collection date between December 2019 and March 23^rd^, 2020 (N = 2,856). We used Nextclade version 0.3.5 (https://clades.nextstrain.org) to infer genomic mutations and filtered for sequences that contained C15324T. This procedure allowed avoidance of homoplasic mutations at this site. Further, we downloaded metadata for all high quality and complete genomes (as of August 12^th^, 2020) irrespective of clade to calculate summary statistics of number of genomes sequenced per country.

### Identification of S-gene D614G mutation in Basel sequences

We screened the early phase Basel sequences for the mutation at nucleotide position 23,403 based on the alignment to the ***Wuhan-Hu-1*** reference sequence MN908947.3. Viral load (Ct-value) of patients that carried lineages with a mutated S-D614G gene (N = 274) were compared to patients that carried the ancestral allele (N = 12) using a Mann-Whitney U test.

## Data Availability

Sequencing data (viral reads only) was submitted to European Nucleotide Archive (ENA) under accession number PRJEB39887, consensus sequences were submitted to GISAID, bioinformatic pipelines are accessible on Github.

https://www.gisaid.org/

https://www.ebi.ac.uk/ena/browser/home

## Ethics

The study was conducted according to good laboratory practice and in accordance with the Declaration of Helsinki and national and institutional standards and was approved by the ethical committee (EKNZ 2020-00769). The clinical trial accession number is NCT04351503 (clinicaltrials.gov).

## Acknowledgements

We thank Daniel Gander, Christine Kiessling, Magdalena Schneider, Elisabeth Schultheiss, Clarisse Straub, and Rosa-Maria Vesco (University Hospital Basel) for excellent technical assistance with sequencing. Calculations were performed at sciCORE (http://scicore.unibas.ch/) scientific computing center at University of Basel, the support from the sciCORE team for the analysis is greatly appreciated. Support for the creation of schematic figures (S2) was provided by BioRender.com. We thank all authors who have shared their genomic data on GISAID especially the Stadler Lab from ETHZ for sharing other Swiss sequences. A full table outlining the originating and submitting labs is included as a supplementary file. No dedicated funding was used for this work.

## Authors contributions

AE and HH devised the project. KL and AG collected and prepared samples and associated data. MS performed the phylogenetic analysis and interpretation, and led the writing and revising of the report. AM constructed the COVGAP bioinformatic pipeline and released it on Github. TR and HSS prepared viral RNA for sequencing, directed the phylogenetic analysis and deposited genomic data to GISAID and ENA. MSch and KKS collected clinical and epidemiological data. MyB and RSS provided geographical expertise. JB, STS and SF provided public health and epidemiological expertise. HP, MSi, CHN, RB, MO, SB, and MB provided clinical expertise and valuable discussion on the results.

All authors commented on the draft report and contributed to the final version.

