## SupplementaRY material for "SARS-CoV-2 outbreak in a tri-national urban area is dominated by a B.1 lineage variant linked to mass gathering events"

Adrian Egli, MD PhD

University Hospital Basel

Petersgraben 4

4031 Basel, Switzerland

### Supplementary Results

#### Basel samples in phylogenetic global context continued

*Cluster B.1.5*

Isolates that are assigned to lineage B.1.5 make up 2.6% of USB isolates (**Figure S6B**). They all share the A20268G mutation. Three unresolved branches defined by at least one additional mutation each, diverge from the internal node consisting of, from top to bottom, six (C25658T), six (C28854T), and one (G25483A, C4893T, C23380A, C26509T [mutations in order of temporal appearance]) Basel area isolates. Individual isolates can exhibit one to three additional mutations. Isolates date from March 13$^{th}$ to March 23$^{rd}$ with an inferred node age of February 19$^{th}$ (CI: January 13$^{th}$-Ferbuary 20$^{th}$, 2020). No social connections for transmission patterns within each branch could be inferred from the available patient data. Searching the clade defining mutations in the nextstrain.org phylogeny we gain the following insights. Mutation C25658T (plus the clade defining A20268G) is found in one isolate (Oman/RESP-20-6701/2020 from March 28$^{th}$); C28854T is found 17 isolates, two of which show no additional mutations (Norway/2088/2020 from March 17$^{th}$, Latvia/045/2020 from March 22$^{nd}$) just like two of our isolates (42193056, 42189239). Derived isolates originate from Switzerland, Scotland, Romania, USA, Taiwan, and England. Mutation G25483A recorded in a single isolate (42202280) is not currently reported in the nextstrain.org phylogeny.

*Cluster B.1.8*

Isolates that are assigned to lineage B.1.8 make up 0.7% of USB isolates (**Figure S6C**). They all share the A24862G mutation. Isolates date from March 14$^{th}$ to March 22$^{nd}$ with an inferred internal node age of February 1$^{st}$ (CI: January 12$^{th}$-March 8$^{th}$, 2020). Two isolates (42191012, 42202147) exhibit the identical mutational pattern (additional T658C, C28829T) but have no known epidemiological link. Our own global comparison identified an isolate from Germany (Germany/NRW-34/2020 from March 16$^{th}$) that exhibits the same mutations. Searching the clade defining mutation in the nextstrain.org phylogeny does not yield better insights into the evolution of the lineages as no isolates with the same pattern could be identified.

*Family clusters within lineage B.2*

We identified eight genomes that were assigned to lineage B.2 (**Figure S6D**). They all share the G26144T mutation that translates into amino acid change ORF3a-G251V and date from March 13$^{th}$ to March 22$^{nd}$ with an inferred internal node age of January 15$^{th}$ (CI: January 13$^{th}$-January 18$^{th}$, 2020). This cluster harbours two household transmission clusters: *Family 2* with two members and *Family 3* with three members. These two clusters share C14805T (synonymous in *ORF1ab*) and exhibit unique additional mutations C9319T (synonymous in *ORF1ab*) and G12278T (ORF1ab-A4005S), G26730T (M-V70F), G29414T (N-A381S), respectively. We find no evidence of further community transmission. These mutational combinations are not currently represented in the full global phylogeny (nextstrain.org), suggesting that quarantine measures were effective in these cases and inhibited further transmission events.

### Supplementary Tables

**Table S1. Counts and description of the in silico mutated genome community used for COVGAP validation**. Each observation consists of the genome position multiplied by the number of samples in which it appears. Attached as additional file.

**Table S2. Nucleotide position in relation to the Wuhan-Hu1 reference sequence that were masked for phylogenetic inferences, due to homoplasies.** Inferred by contributors to https://github.com/W-L/ProblematicSites_SARS-CoV2.

| Start position | End position |
|---|---|
| 635 | 635 |
| 2091 | 2091 |
| 2094 | 2094 |
| 3145 | 3145 |
| 3564 | 3564 |
| 4050 | 4050 |
| 5736 | 5736 |
| 6869 | 6869 |
| 8022 | 8022 |
| 8790 | 8790 |
| 10129 | 10129 |
| 11074 | 11074 |
| 11083 | 11083 |
| 11535 | 11535 |
| 13402 | 13402 |
| 13408 | 13408 |
| 13476 | 13476 |
| 13571 | 13571 |
| 14277 | 14277 |
| 15922 | 15922 |
| 16887 | 16887 |
| 19484 | 19484 |
| 21575 | 21575 |
| 22335 | 22335 |
| 24389 | 24389 |
| 24390 | 24390 |
| 24933 | 24933 |
| 26549 | 26549 |
| 29037 | 29037 |
| 29553 | 29553 |

**Table S3. List of countries that recorded genomes with mutation C15324T and number of total genomes sequenced until March 23rd 2020.**

| Country | number genomes with C15324T | Total genomes sequenced until March 23rd | % genomes with mutation | % of population sequenced | Population |
|---|---|---|---|---|---|
| **Argentina** | 1 | 4 | 25.00 | 0.00001 | 45,195,774 |
| **Australia** | 14 | 1092 | 1.28 | 0.00428 | 25,499,884 |
| **Austria** | 3 | 244 | 1.23 | 0.00271 | 9,006,398 |
| **Belgium** | 40 | 268 | 14.93 | 0.00231 | 11,589,623 |
| **Benin** | 1 | 6 | 16.67 | 0.00005 | 12,123,200 |
| **Bosnia and Herzegovina** | 2 | 12 | 16.67 | 0.00037 | 3,280,819 |
| **Brazil** | 1 | 226 | 0.44 | 0.00011 | 212,559,417 |
| **Canada** | 7 | 405 | 1.73 | 0.00107 | 37,742,154 |
| **Chile** | 2 | 120 | 1.67 | 0.00063 | 19,116,201 |
| **Costa Rica** | 1 | 40 | 2.50 | 0.00079 | 5,094,118 |
| **Democratic Republic of the Congo** | 11 | 35 | 31.43 | 0.00004 | 89,561,403 |
| **England** | 4 | 5643 (UK) | 0.01 | 0.00831 | 67,886,011 |
| **France** | 69 | 369 | 18.70 | 0.00057 | 65,273,511 |
| **Germany** | 2 | 147 | 1.36 | 0.00018 | 83,783,942 |
| **Hungary** | 2 | 18 | 11.11 | 0.00019 | 9,660,351 |
| **Iceland** | 5 | 522 | 0.96 | 0.15297 | 341,243 |
| **India** | 1 | 119 | 0.84 | 0.00001 | 1,380,004,385 |
| **Israel** | 1 | 72 | 1.39 | 0.00083 | 8,655,535 |
| **Japan** | 3 | 343 | 0.87 | 0.00027 | 126,476,461 |
| **Luxembourg** | 24 | 116 | 20.69 | 0.01853 | 625,978 |
| **Morocco** | 3 | 13 | 23.08 | 0.00004 | 36,910,560 |
| **Netherlands** | 2 | 617 | 0.32 | 0.00360 | 17,134,872 |
| **Oman** | 1 | 21 | 4.76 | 0.00041 | 5,106,626 |
| **Portugal** | 8 | 570 | 1.40 | 0.00559 | 10,196,709 |
| **Russia** | 1 | 59 | 1.69 | 0.00004 | 145,934,462 |
| **Scotland** | 4 | 5643 (UK) | 0.01 | 0.00831 | 67,886,011 |
| **Senegal** | 3 | 24 | 12.50 | 0.00014 | 16,743,927 |
| **South Korea** | 1 | 196 | 0.51 | 0.00038 | 51,269,185 |
| **Switzerland** | 57 (386)* | 213 (675)* | 26.8 (57.2)* | 0.00780 | 8,654,622 |
| **Taiwan** | 3 | 95 | 3.16 | 0.00040 | 23,816,775 |
| **USA** | 1 | 4150 | 0.02 | 0.00125 | 331,002,651 |
| **Vietnam** | 1 | 59 | 1.69 | 0.00006 | 97,338,579 |

**** Number in brackets summarize counts of genomes from GISAID plus genomes from this study***

**Table S4. GISAID identifiers and dates of sampling for all sequences that belong to emerging clade 20A/15324T with a collection date until March 23rd, 2020 (N = 279). Supplied as additional file.**

**Table S5. Diversity indices for SARS-CoV-2 lineages in Switzerland and neighbouring countries.**

| Country | Coefficient of co-variation | Shannon Entropy H’ | Shannon Diversity | Simpson Concentration Index D’ | Simpson Diversity |
|---|---|---|---|---|---|
| **Austria** | 1.577 | 1.680 | 5.365 | 0.2679 | 3.7322 |
| **France** | 1.762 | 0.421 | 1.524 | 0.8144 | 1.2278 |
| **Germany** | 1.358 | 1.637 | 5.137 | 0.2583 | 3.8715 |
| **Italy** | 0.751 | 1.067 | 2.908 | 0.3971 | 2.5181 |
| **Switzerland** | 2.768 | 0.869 | 2.385 | 0.6156 | 1.6243 |

### Supplementary Figures

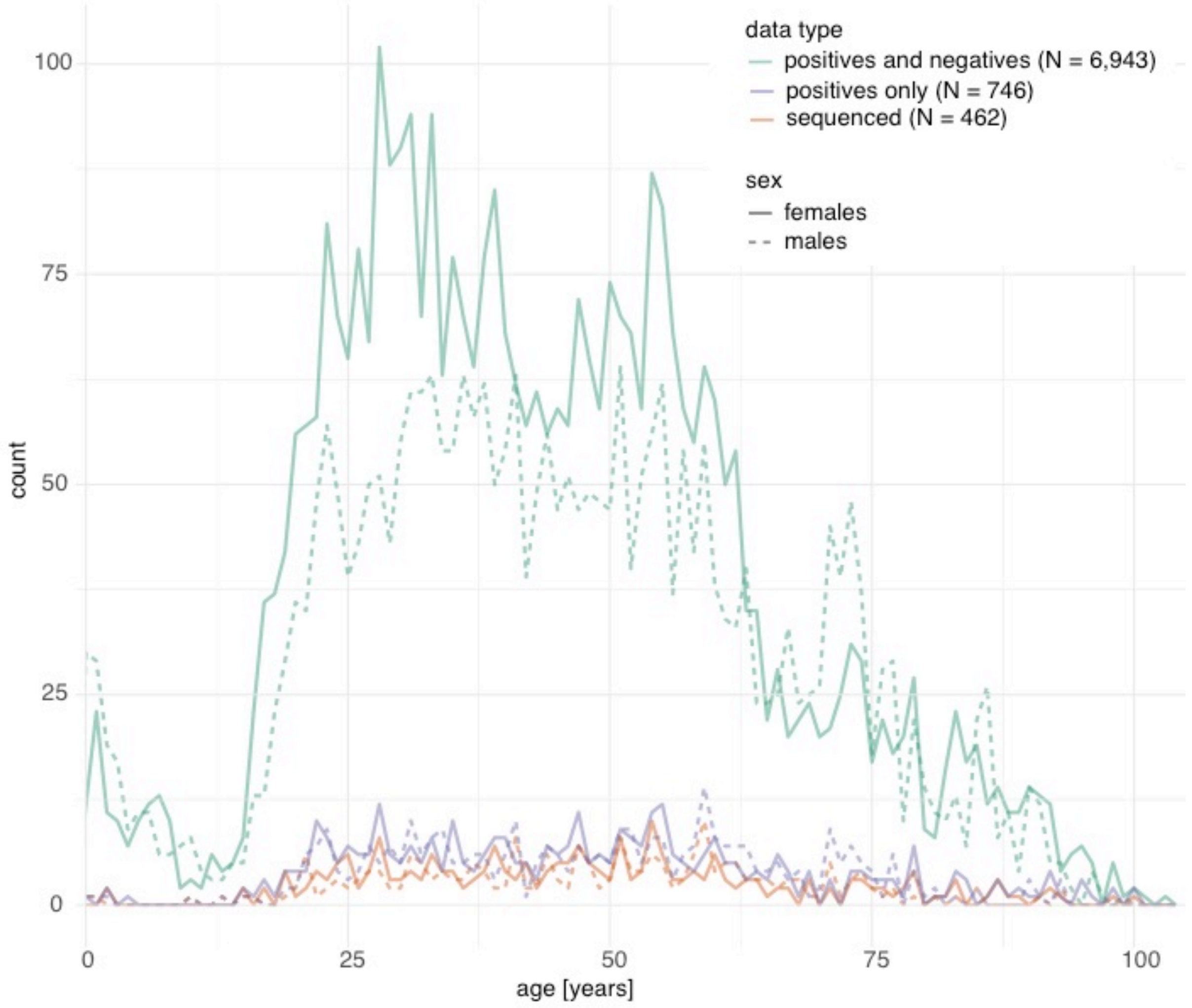

**Figure S1. Age distribution by sex for the time period between February 24th and March 23rd for all tests, positive tests, and for patient isolates from which whole genomes were generated.** Solid lines for females, dashed lines for males.

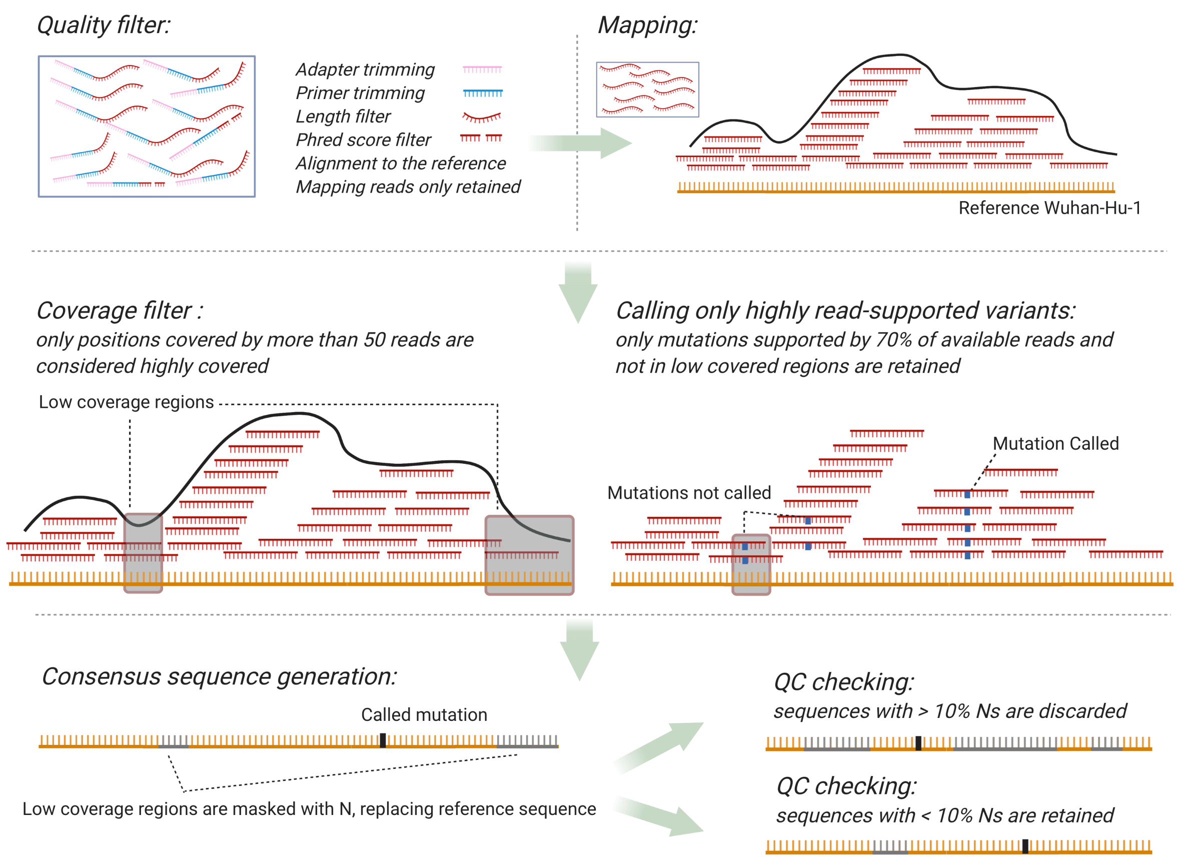

**Figure S2. The COVGAP pipeline.** The steps shown ensure the calculation of high quality consensus sequences. Particularly, information on read coverage is retained and used both in the variant calling procedure and in the draft of the consensus independently from the called variants. Finally, the quality of the genome from each sample is scored by %Ns, which determines whether the produced sequence is retained or discarded.

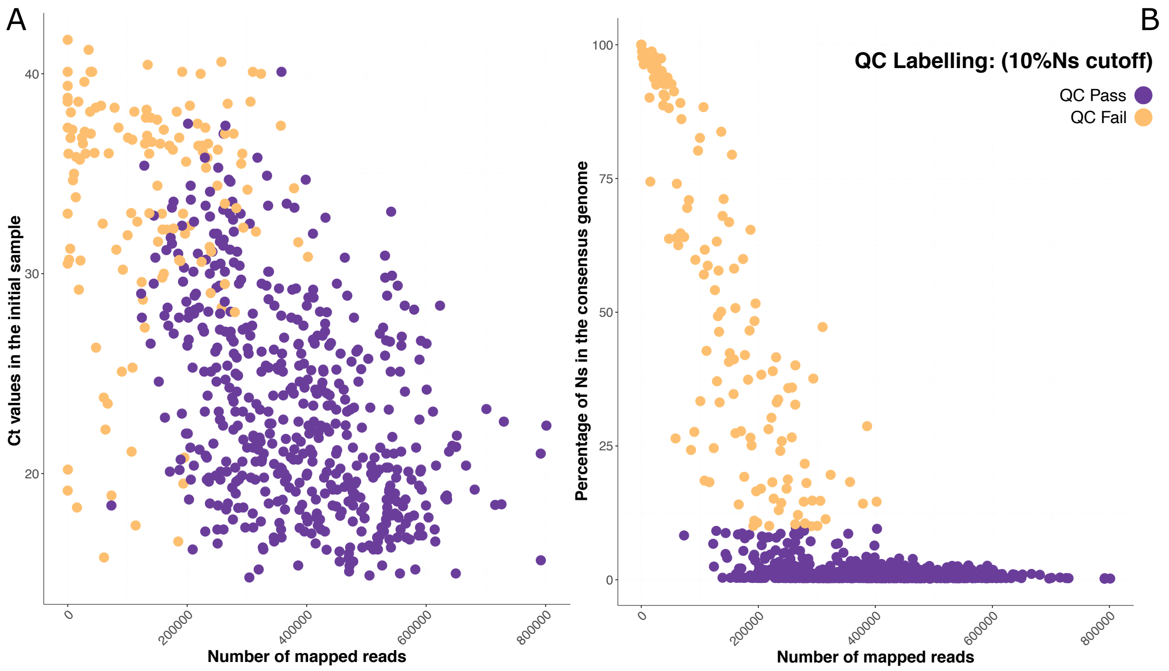

**Figure S3. COVGAP evaluation of sequencing quality parameters.**

Of the original 746 samples, 689 successfully sequenced. Number of mapped reads across all SARS-CoV-2 positive samples successfully sequenced from 26th of February till 23th of March: (n=689), of which 533 passed the quality filter, and 156 failed. 468 of the samples passing the quality filters were matching the cohort eligibility criteria and therefore were further described in the present study. **A.** Number of mapped reads against Ct values from diagnostic tests; **B.** Number of mapped reads against percentage of Ns in the consensus.

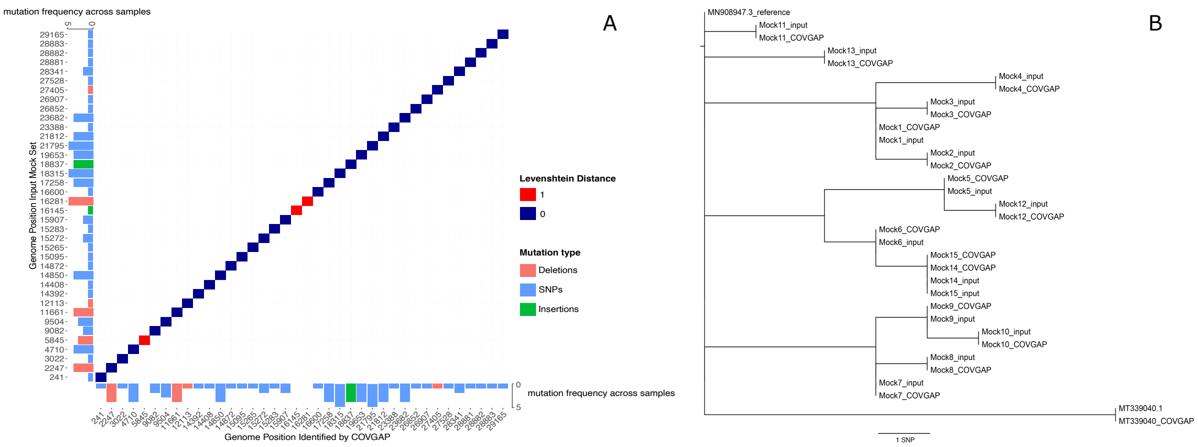

**Figure S4. COVGAP identifies all SNPs in the mock genomes. A**. Levenshtein distance between the mutations in the input mock genomes (y axis) and in the genomes recovered by COVGAP (x axis); the marginal plots show the frequency of presence / detection across samples. Only two deletions (5845, 16281) and one insertion (16145) were not detected. **B**. Phylogeny of input and COVGAP-derived consensus output genomes, showing that all SNPs were identified.

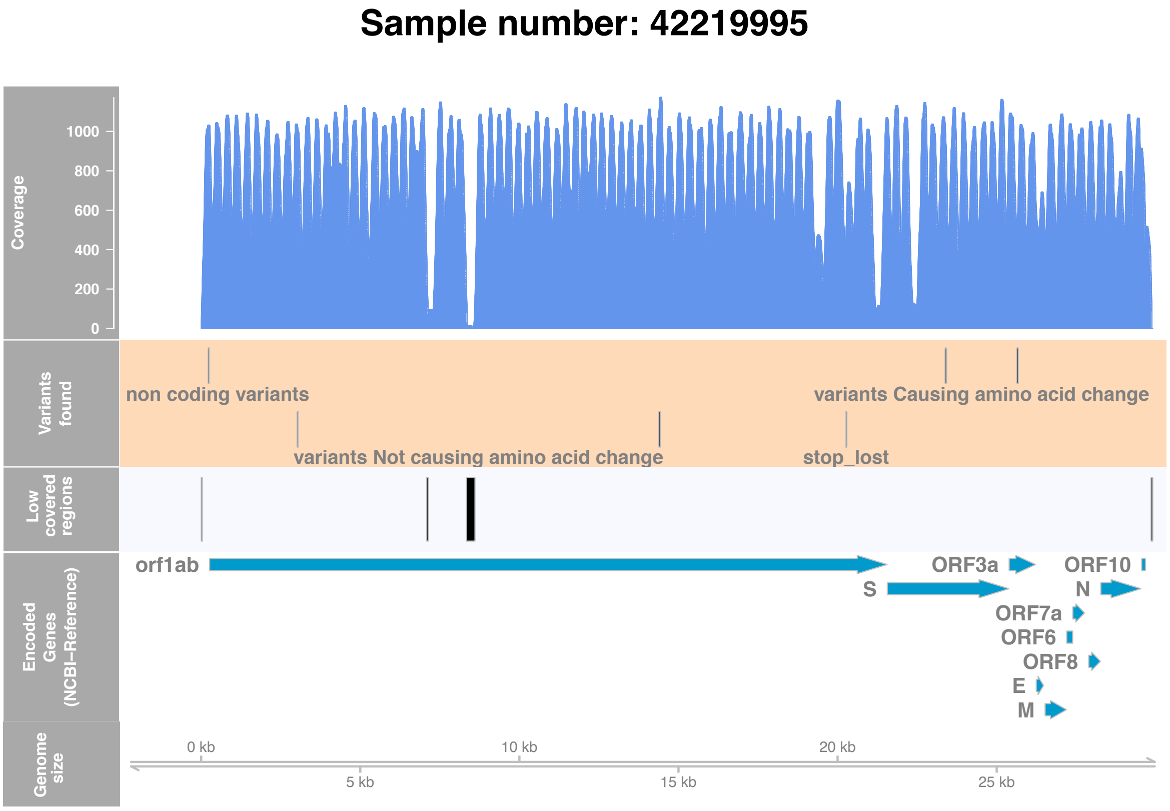

**Figure S5. Representative diagnostic output from COVGAP.** This output generated per sample, indicates (from top to bottom): **A**. the coverage –not represented if over 1000x; **B**. which variants were detected in which position of the genome, and their corresponding annotation; **C**. low coverage regions (under 50x); **D**. genome annotation; and **E**. genome size markers as reference. Of note, a report generated in parallel provides further information on the variants, including which amino acids are affected by the variant.

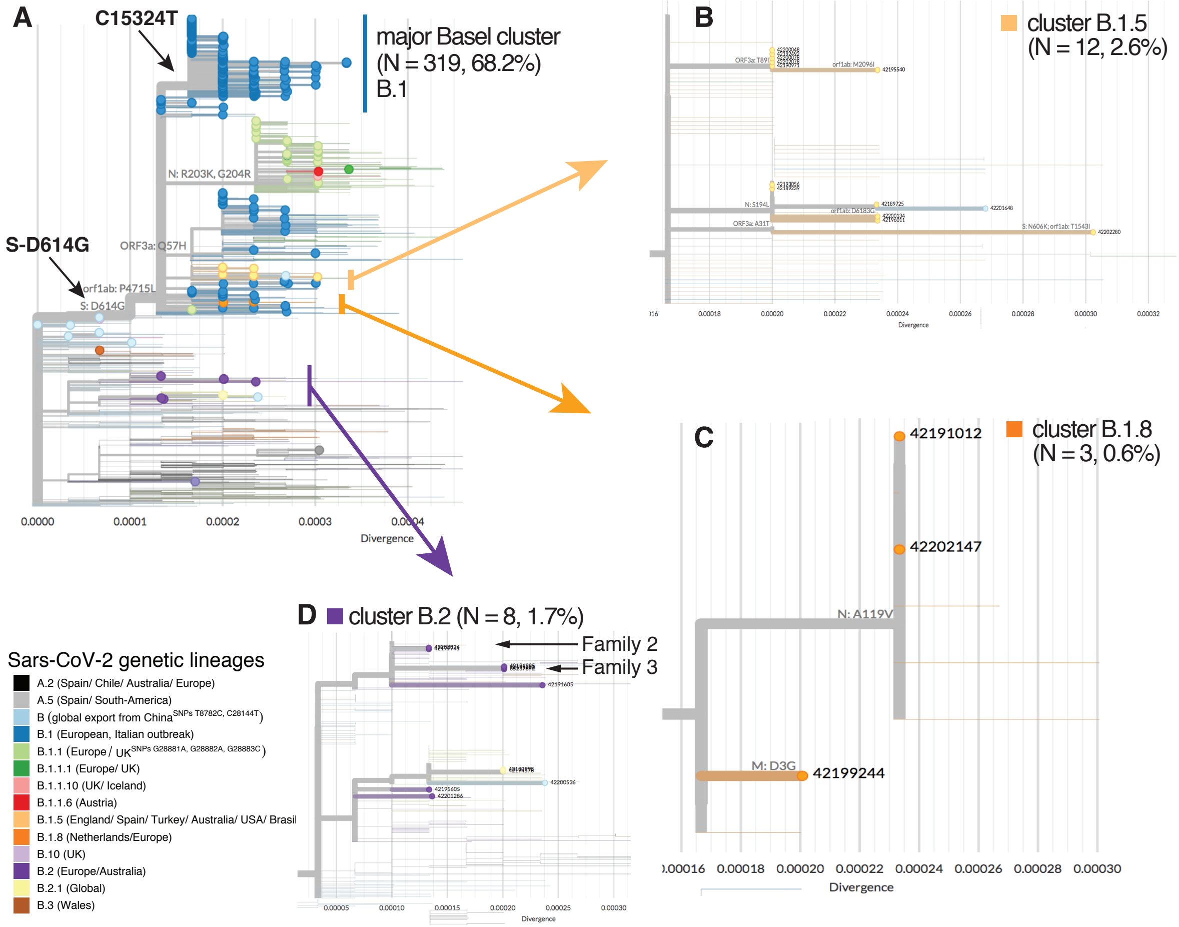

**Figure S6. Divergence tree and zoom into additional sequence clusters, which did not result in large community spread. A**. Isolates from Basel area cohort in global context. **B**. A small clade assigned to B.1.5 consists of two clusters with an accumulation of samples from Basel. **C**. Cluster within lineage B.1.8 with two Basel samples without known epidemiological link. **D**. Two family cluster within lineage B.2 that did not spread further into the Basel community.
